# Depression predicts chronic pain interference in racially-diverse, low-income patients

**DOI:** 10.1101/2021.06.17.21259108

**Authors:** Benjamin C. Nephew, Angela C. Incollingo Rodriguez, Veronica Melican, Justin J. Polcari, Kathryn E. Nippert, Mikhail Rashkovskii, Lilly-Beth Linnell, Ruofan Hu, Carolina Ruiz, Jean A. King, Paula Gardiner

## Abstract

**Background:** Chronic pain is one of the most common reasons adults seek medical care in the US, with estimates of prevalence ranging from 11% to 40%. Mindfulness meditation has been associated with significant improvements in pain, depression, physical and mental health, sleep, and overall quality of life. Group medical visits are increasingly common and are effective at treating myriad illnesses including chronic pain. Integrative Medical Group Visits (IMGV) combine mindfulness techniques, evidence based integrative medicine, and medical group visits and can be used as adjuncts to medications, particularly in diverse underserved populations with limited access to non-pharmacological therapies.

**Objective and Design:** The objective of the present study was to use a blended analytical approach of machine learning and regression analyses to evaluate the potential relationship between depression and chronic pain in data from a randomized clinical trial of IMGV in socially diverse, low income patients suffering from chronic pain and depression.

**Methods:** This approach used machine learning to assess the predictive relationship between depression and pain and identify and select key mediators, which were then assessed with regression analyses. It was hypothesized that depression would predict the pain outcomes of average pain, pain severity, and pain interference.

**Results:** Our analyses identified and characterized a predictive relationship between depression and chronic pain interference. This prediction was mediated by high perceived stress, low pain self-efficacy, and poor sleep quality, potential targets for attenuating the adverse effects of depression on functional outcomes.

**Conclusions:** In the context of the associated clinical trial and similar interventions, these insights may inform future treatment optimization, targeting, and application efforts in racially diverse, low income populations, demographics often neglected in studies of chronic pain.

## INTRODUCTION

Chronic pain is one of the most common reasons adults seek medical care in the United States (U.S.), with estimates of chronic pain prevalence ranging from 11% to 40% [1]. In turn, patients with chronic pain are more likely to use health care services than the general population, which increases associated medical costs. Chronic pain has also been associated with overall poor health and other comorbidities, such as depression, stress, and insomnia, with particularly high prevalence in patients already prone to healthcare disparities [2]. Previous research indicates that residency in low-income neighborhoods is associated with higher cost use of the health care system. Accordingly, addressing social determinants of health has been proffered as a critical step in improving overall health and reducing costs, especially with regards to chronic pain [3]. Additionally, findings from a 12-year longitudinal study highlight the importance of considering socioeconomic status when assessing chronic pain in individuals [4]. This study identified large magnitude differences in chronic pain scores, with low income ethnic minorities experiencing greater pain compared to more wealthy non-minority individuals [4]. Furthermore, how this chronic pain is managed often differs between income groups, as lower income patients are more likely to take prescription pain medications and less likely to use exercise to alleviate chronic pain than higher income patients [5].

There is substantial evidence of links between chronic pain and depression, as 30-60% of patients with chronic pain experience depression [6]. This is not surprising given the shared neurobiological (e.g., genes, hormones, brain networks) and behavioral (e.g., sleep disturbance) factors across both conditions [7]. Exposure to psychosocial stress has been specifically implicated in depression in pain patients [8], and comorbid chronic pain and depression has been associated with poorer prognosis and greater disability compared to patients with chronic pain alone [6]. Furthermore, prescription medication interventions for depression, including serotonin norepinephrine reuptake inhibitors (SNRIs) and tricyclic antidepressants, are effective at reducing the symptoms of both depression and chronic pain [6]. Clinical observations support theories of a reciprocal interaction between chronic pain and depression, where depression symptoms worsen pain perception and chronic pain accentuates depression behaviors such as poor sleep [9]. This is likely mediated by psychosocial stress in low SES populations.

The high prevalence and treatment resistance of chronic pain, combined with the adverse consequences of pain medication dependence, have stimulated increased interest in mindfulness-based interventions (Hilton, Hempel et al. 2017). Common interventions applied for chronic pain reduction include mindfulness-based stress reduction (MBSR), mindfulness-based cognitive therapy (MBCT), and integrative treatments that combine mindfulness techniques with other conventional and complementary medicines. Mindfulness elements are thought to allow individuals to reframe experiences such as chronic pain by refocusing the mind on the present and increasing awareness of external surroundings and inner sensations (Grant and Rainville, 2009).

Mindfulness meditation is associated with statistically significant improvements in depression, physical and mental health, pain self-efficacy, and sleep [10]. However, the magnitude of health improvement can vary across studies. Cross-sectional cohort studies of people engaging in long-term meditation practices find that higher levels of mindfulness are associated with lower pain intensity ratings and changes in the structure of brain regions related to pain perception (Luders, Phillips et al. 2012). However, studies of mindfulness interventions as a treatment for chronic pain have heterogeneous results overall. While some studies focus on specific types of pain such as low back pain (Cramer, Haller et al. 2012), fibromyalgia (Kozasa, Tanaka et al. 2012) or chronic migraines, others broadly target chronic pain. Recent systematic reviews (Hilton, Hempel et al. 2017, Majeed, Ali et al. 2018) conclude that treatment of chronic pain with mindfulness interventions has moderate success, and more high-quality research is needed to support a recommendation for the use of mindfulness meditation for any chronic pain symptomology.

Group medical visits are an increasingly common intervention and are effective at treating myriad illnesses, including chronic pain. Within these group visits, peer support from group members, an understanding of self-healing, and overall relationships are built with both the clinician and other participants [11]. These experiences suggest that group medical visits can be a viable intervention for alleviating chronic pain. Our prior work evaluating 65 patients with chronic pain during group medical visits over a 12-month course reported decreases in pain level, perceived stress, and depression as well as improvements in sleep quality [12]. A 2017 study of a Mindfulness Based Stress Reduction program for 20 low-income minority adults with chronic pain and comorbid depression determined that group treatment contributed to enhanced coping with chronic pain and improvement in the sense of control over one’s health condition [13]. Integrative medical group visits (IMGV), which focus on increasing access to integrative health care, have also been particularly effective in managing and treating chronic conditions [14]. They can be used as adjuncts to medications, particularly in underserved diverse, low-income populations with limited access to non-pharmacological therapies. There are benefits to both the patients and clinicians participating in integrative group medical visits, with improvements in patient physical and mental health [14].

Sleep disorders are also extremely prevalent in patients with chronic pain, and depression and anxiety frequently develop alongside insomnia (Wilson, Eriksson et al. 2002). These relationships are often bidirectional, such that pain leads to sleep disturbances and patients with a history of insomnia are more likely to develop chronic pain (Keilani, Crevenna et al. 2018), and sleep quality may mediate a variety of specific pain outcomes. Patients with major depressive disorder and concurrent insomnia report the highest levels of pain outcomes, including affective distress (Wilson, Eriksson et al. 2002), and increased levels of pain were observed in patients with insomnia even in the absence of major depression. Another potential mediator of chronic pain is exposure to chronic stress.

Chronic stress is recognized as a primary cause of disability and mortality worldwide and is often associated with chronic pain (Davis, Holmes et al. 2017). Pain can often result in increased levels of perceived stress which can then lead to more pain in a bidirectional relationship, similar to covariation between sleep and pain (Lunde and Sieberg, 2020). Moreover, stress not only triggers pain but also modulates the perception of pain, where the effects of psychological state on pain perception impacts the success of treatments. For example, when recovery does not meet a person’s expectations, this may result in negative thoughts and an enhanced perception of pain (Linton and Shaw, 2011). These findings highlight the importance of the application of complementary medicine in pain management. These treatments can safely reduce the adverse impacts of chronic pain through stress reduction. For example, mind-body interventions provide individuals with the tools to practice relaxation, reduce stress, and attenuate pain perception.

Our recent randomized control trial (RCT) assessed the effectiveness of IMGV compared to a Primary Care Provider (PCP) visit in patients with chronic pain and depression in a 9-week single-blind two arm randomized control trial with a 12-week maintenance phase (intervention-medical groups; control-primary care provider visit) [15]. All participants (N=155) were randomized (1:1) to either intervention (IMGV) or control group. There were no group differences in average pain level in predominantly low income racially diverse adults with nonspecific chronic pain and depressive symptoms. However, the IMGV group had fewer emergency department visits at 9 weeks, and this group reported reduced pain medication use at 21 weeks compared to controls. These results demonstrate that low-income racially diverse patients will attend medical group visits that focus on non-pharmacological techniques.

The objective of the present study was to use a blended analytical approach of machine learning and hierarchical linear regression analyses to evaluate the potential relationship between depression and pain in the data from our IMGV RCT in a racially diverse low-income population. This approach used machine learning to evaluate the predictive relationship between depression and pain measures and identify and select key mediators, which were then assessed with linear regression. It was hypothesized that depression would predict the pain outcomes of average pain, pain severity, and pain interference.

## METHODS

### Study Setting

The study was approved by the Boston University Medical Campus Institutional Review Board (IRB) and the community health center’s (CHC) research committees (IRB Approval Number: H33096). We registered this randomized controlled trial (RCT) in the international trial register [ClinicalTrials.gov: Identifier NCT02262377]. The RCT was conducted at an ambulatory primary care clinic in the outpatient building at Boston Medical Center (BMC) in Boston MA and at two community health centers: Codman Square Health Center (CSHC) and Dorchester House Health Center (DHHC). BMC and the two affiliated federally qualified health centers serve low income, racially and ethnically diverse populations with health disparities in the treatment of chronic pain and depression. BMC is a private, not-for-profit, academic medical center with 567 beds. DHHC is a 3-story modern health facility with 40 clinical exam rooms. DHHC has approximately 30,000 active patients. CSHC has 22 exam rooms, and three new rooms for group medical visits. CSHC has approximately 21,000 active patients. These locations were chosen because they are the setting where there is a high prevalence of low-income diverse patients with chronic pain and depression who do not have access to non-pharmacological treatments. All three sites were accessible by public transportation.

### IMGV Intervention

The IMGV intervention includes three concurrent deliveries of the same self-management curriculum delivered with different formats–an in-person MGV, and two adjunct companion technologies available on a computer tablet provided to the intervention participant. The first technology was the Our Whole Lives (OWL), an e-Health toolkit platform, and the second technology was an Embodied Conversational Agent (ECA). A detailed description of the IMGV self-management intervention has previously been described [34]. The IMGV consists of a total of ten in-person medical group visits each lasting 2.5-hours conducted weekly from week 1 to week 9 (9 in-person sessions plus OWL/ECA). This is followed by a 12-week maintenance phase where there is access to the technology only (OWL/ECA). A tenth and final in-person session is conducted at week 21.

At the beginning of each session of the IMGV, participants measured their vital signs, mood, and pain levels. They then met individually with a trained physician (a co-facilitator) for a medical assessment. A trained non-physician facilitator (see below) then led mindfulness practices adapted from Mindfulness Based Stress Reduction (meditation, body scan, etc). Patients were instructed in the principles of mindfulness and other self-management techniques (such as acupressure and self-massage). Each week, the physician facilitated a discussion on health topics such as stress, insomnia, depression, chronic pain cycle, activity, and healthy food choices. Finally, the IMGV ended with a healthy meal, which mirrored the healthy nutrition topic in each session. In addition to a physician, an experienced co-facilitator with training in mindfulness (certified MBSR instructor, yoga and meditation teacher) attended all groups. Facilitators were mentored via direct observation of two pilot group visits, one-on-one meetings, and phone calls by an experienced MBSR trained faculty.

To reinforce all content delivered in the in-person group, an internet-based platform called Our Whole Lives; an electronic health toolkit (OWL) delivered the same in-person curriculum. OWL could be accessed with a computer, smart phone, or tablet. The ECA, a female automated character, emulated the conversational behavior of an empathic coach [48]. The ECA (Gabby) reviewed all the content discussed in the IMGV with the participants outside of the in-person group. A Dell Venue 8 Pro tablet was distributed to all intervention participants in the first session of the group.

After the nine-week in-person group visit phase concluded, the intervention participants entered a 12-week maintenance phase. The intervention participants retained the study tablet and continued to have access to the ECA and the OWL website. At the end of the 21 weeks, there was one final in-person group visit.

A trained study RA directly observed all groups and assessed the facilitator’s adherence to the intervention components through a monitoring and evaluation checklist. These checklists were used to assess each MGV session at all sites during the study. All participants randomized to the control group were asked to visit their PCP during the study period (baseline to 21 weeks). We verified a PCP clinical visit via electronic medical record (EMR) documentation. We did not collect data on the duration or content of the visit.

### Clinical Outcomes

Outcome data were collected at baseline, 9 weeks, and 21 weeks. Baseline demographic data included: age, gender, race, ethnicity, income, work status, and education. Our primary outcomes included in the present analysis consist of: 1) self-reported pain measured by the Brief Pain Inventory [(BPI) pain interference, pain severity, and average pain score in the last seven days] [16] and 2) depression level measured by the PHQ-9, a self-reported depression screen [17]. BPI pain interference, pain severity, and average pain are on a 0-10-point scale. The higher the score, the more severe the pain. PHQ-9 is on a 0-27-point scale. The higher the score, the more severe the depression.

Secondary outcomes included were perceived stress, pain self-efficacy, and sleep quality. The Perceived Stress Scale (PSS) is a self-reported questionnaire designed to assess “the degree to which individuals appraise situations in their lives as stressful [18].” Pain self-efficacy was measured with the Pain Self-Efficacy Scale (PSEQ) and ranged from 0–60 points. High PSEQ scores are associated with higher confidence to function with pain [19]. Sleep quality was measured using the Pittsburgh Sleep Quality Index (PSQI) which assesses sleep quality and disturbances over a 1-month time interval (Buysse, 1989). Seven component scores including subjective sleep quality, sleep latency, sleep duration, habitual sleep efficiency, sleep disturbances, use of sleeping medication, and daytime dysfunction are used to generate one global score (Buysse, 1989).

### Machine Learning

Lasso regression and random forest regressor models were run to determine which features were most predictive of pain severity and interference levels. These models were selected because they provide information about the relative importance of the independent variable in predicting the dependent variable. The dependent variables used in the models were the Brief Pain Inventory (BPI) severity and interference sub-scores and the 0 to 10 average pain score. Each dependent variable was evaluated at 9 and 21 weeks and were run with independent variables from baseline. Models were run using control, intervention, and both groups combined with the complete clinical data set. Independent variables that were missing more than 60% of values were dropped, and the remaining variables’ missing values were imputed with the mean. The data were then transformed using the MinMaxScaler function in the Python scikit-learn library, which scaled each variable between zero and one. The rankings of the top four features were combined across the three dependent variables to summarize which features had the highest relative importance in predicting pain, which were depression, perceived stress, pain self-efficacy, and sleep quality.

### Regression Analyses

A series of separate hierarchical linear regression analyses tested various psychosocial constructs measured at 9 weeks (pain self-efficacy, perceived stress, and sleep quality) as mediators of the relationship between baseline depression and pain outcomes at 21 weeks (average pain, pain severity, and pain interference).

## RESULTS

Three hundred forty-three patients were assessed for eligibility, 209 patients were eligible, and 159 were enrolled and randomized to intervention (80) or control. The consort diagram and baseline characteristics were previously published [15]. Of the 155 participants, the average age was 51 years old, 86% identified as female, 56% identified as black, 36% identified as “other” race, and 19% identified as white. Sixty three percent earned less than $30,000 a year, only 21% worked full or part time, 14% were on unemployed and 42% were on work disability. Common co-morbidities in the participants were hypertension (41%), insomnia (26%), anxiety (28%), Post-Traumatic Stress Disorder (PTSD) (16%), and any substance use disorder (25%). There were no significant differences for baseline characteristics (Table 1). The average PHQ-9 score for depressive symptoms was 12 at baseline, which is characterized as moderate depression.

**Table 1:**
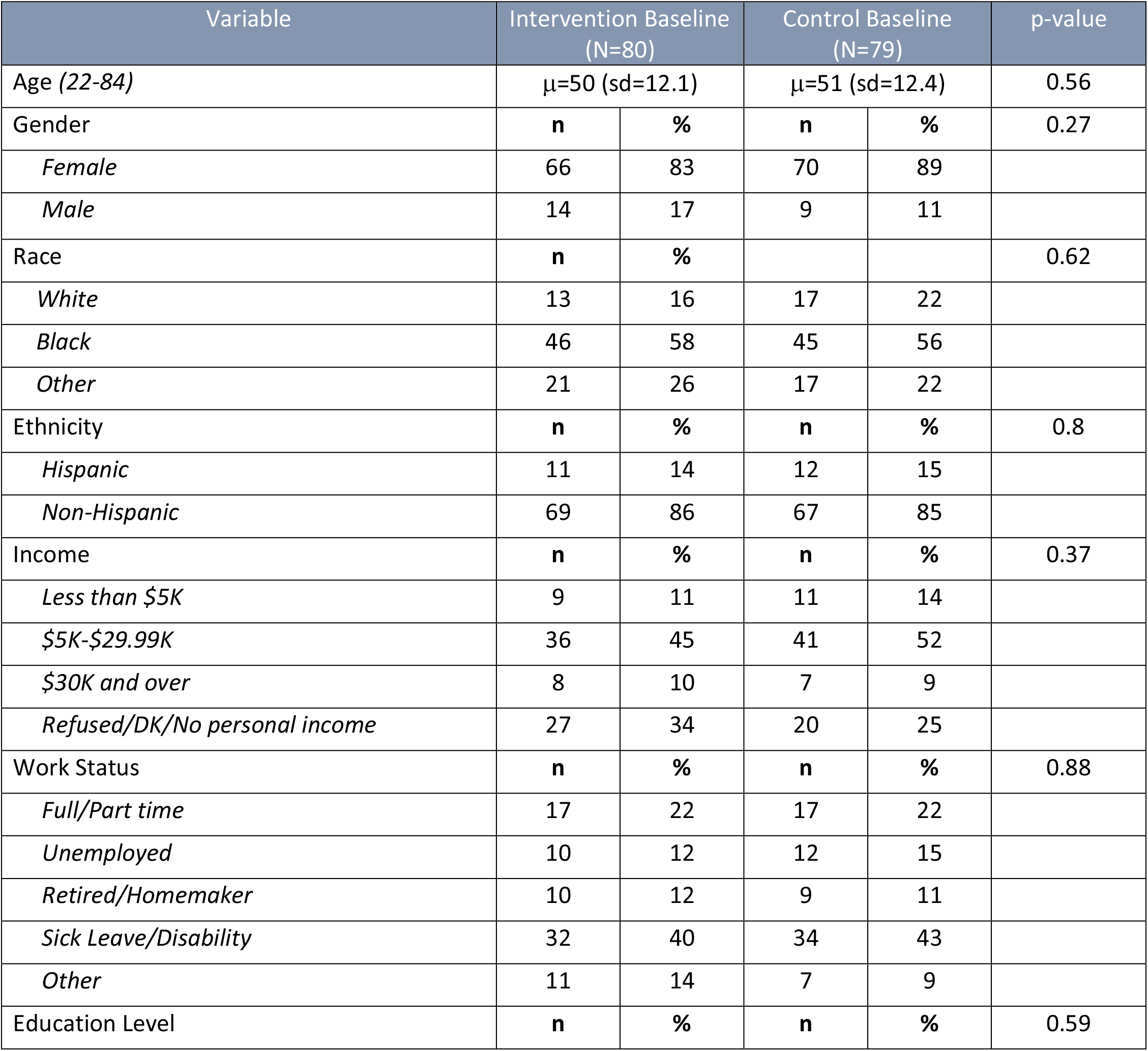
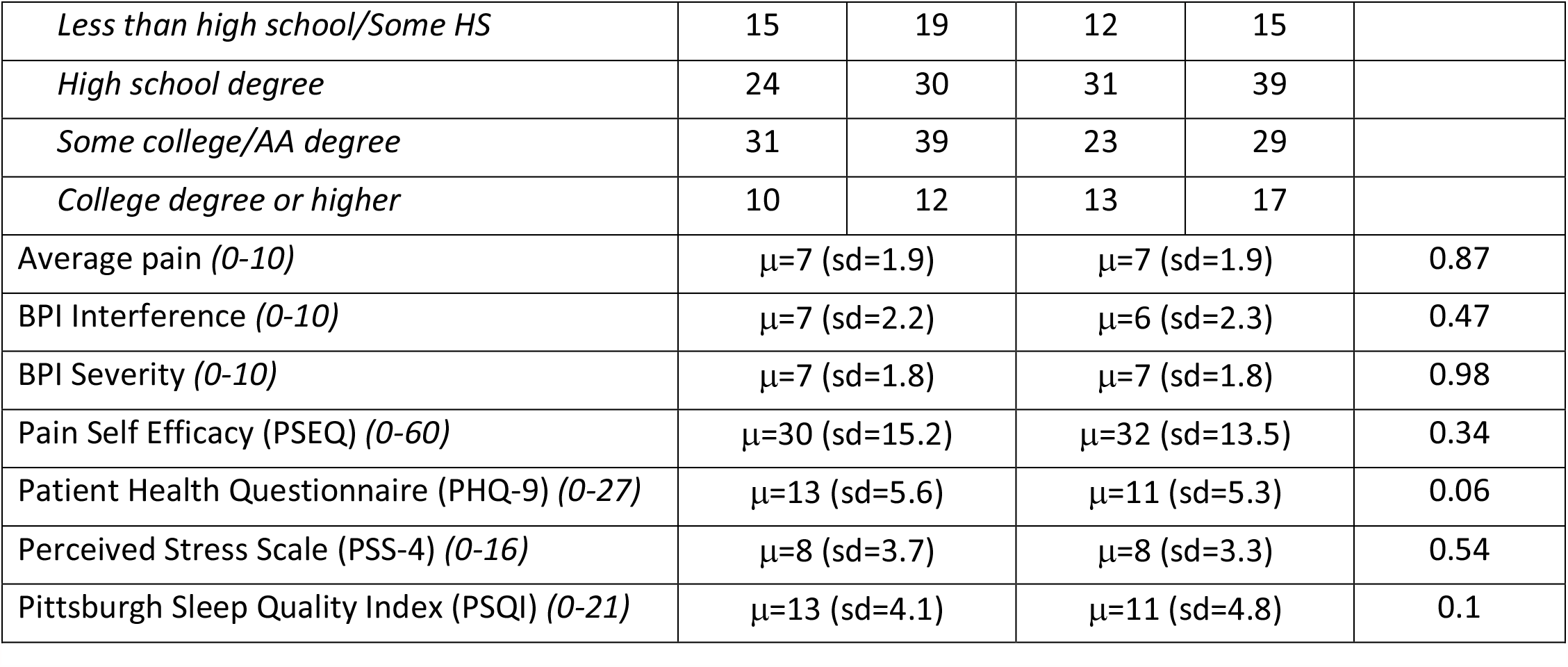
Baseline patient characteristics by treatment group.

### Pain Interference

Baseline depression was a significant predictor of pain interference at 21 weeks, *B* = 0.14, *p* = .001. Therefore, the following variables measured at 9 weeks were tested as potential mediators of this relationship: perceived stress, sleep quality, and pain self-efficacy.

Perceived stress significantly mediated the relationship between baseline depression scores and pain interference at 21 weeks. Specifically, baseline depression significantly and positively predicted perceived stress at 9 weeks, which in turn significantly and positively predicted pain interference at 21 weeks. When accounting for this mediation via perceived stress, baseline depression still uniquely predicted pain interference at 21 weeks.

Sleep quality also significantly mediated the relationship between baseline depression and pain interference at 21 weeks in the same direction as perceived stress.

Pain self-efficacy significantly mediated the relationship between depression and pain interference. Baseline depression significantly and negatively predicted pain self-efficacy at 9 weeks, which in turn significantly and negatively predicted pain interference at 21 weeks. See table 2 for regression coefficients and figure 1 for a summary of the significant findings.

**Table 2:** Hierarchical linear regression analyses testing mediators of depression to interference.

| Perceived Stress (*n* = 134) |  |  |  |  |
| --- | --- | --- | --- | --- |
| Path | *B* | *SE B* | *B* | *p*-value |
| Baseline depression to perceived stress | 0.28 | 0.05 | .43 | **<.001** |
| Perceived stress to interference<br>(accounting for baseline depression) | 0.18 | 0.07 | .24 | **.009** |
| Baseline depression to interference<br>(controlling for perceived stress) | 0.10 | 0.04 | .20 | **.029** |
| Sleep Quality<sup>1</sup> (*n* = 133) |  |  |  |  |
| Path | *B* | *SE B* | *B* | *p*-value |
| Baseline depression to sleep quality | 0.28 | 0.07 | .33 | **<.001** |
| Sleep quality to interference<br>(accounting for baseline depression) | 0.18 | 0.05 | .31 | **<.001** |
| Baseline depression to interference<br>(controlling for sleep quality) | 0.10 | 0.04 | .20 | **.020** |
| Pain Self-Efficacy (*n* = 151) |  |  |  |  |
| Path | *B* | *SE B* | *B* | *p*-value |
| Baseline depression to pain self-efficacy | -0.71 | 0.22 | -.26 | **.002** |
| Pain self-efficacy to interference<br>(accounting for baseline depression) | -0.08 | 0.01 | -.44 | **<.001** |
| Baseline depression to interference<br>(controlling for pain self-efficacy) | 0.09 | 0.04 | .19 | **.017** |

Note. Predictors measured at baseline; mediators at 9 weeks; outcomes at 21 weeks
<sup>1</sup>Higher values indicate worse sleep

**Figure 1:**
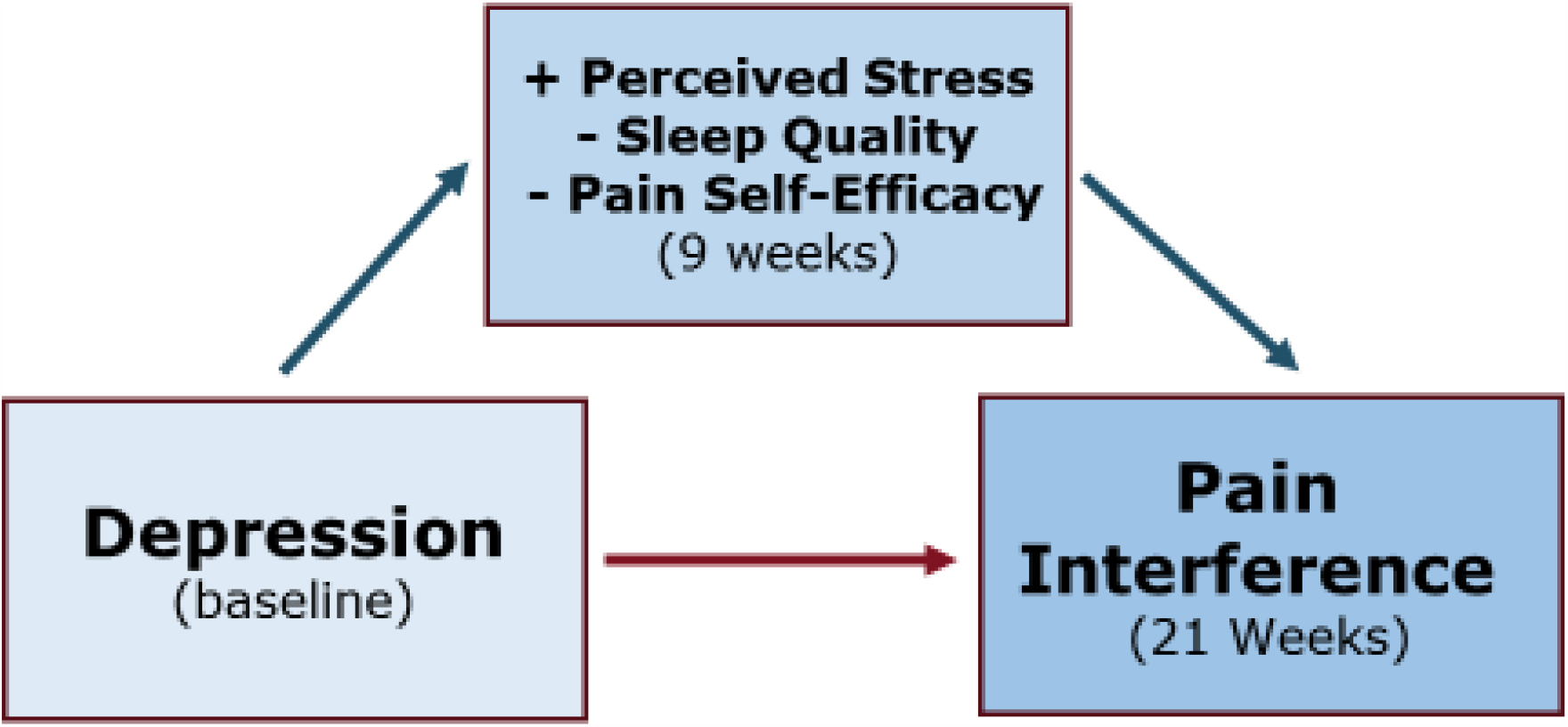
Summary of the mediation of the prediction of pain interference by depression by perceived stress, sleep quality, and pain self-efficacy.

### Average Pain

Baseline depression did not significantly predict average pain, *B* = 0.01, *SE* = 0.03, *p* = .709. Therefore, none of the above psychosocial constructs were tested as mediators.

### Pain Severity

Baseline depression did not significantly predict pain severity as well, *B* = 0.02, *SE* = 0.03, *p* = .601. Again, no mediation analyses were conducted.

## DISCUSSION

The present study identified and characterized a predictive relationship between depression and chronic pain interference in a racially diverse, low-income population. Notably, this sample represents demographic characteristics often neglected in studies of chronic pain. This prediction was mediated by perceived stress, pain self-efficacy, and poor sleep quality, such that depression predicted adverse indicators of these mediators, which in turn predicted higher pain interference. In contrast, depression did not predict reported average pain or pain severity. Increased focus on depression symptomology and the roles of stress, self-efficacy, and sleep in the research and treatment of chronic pain in similar populations may lead to advances in chronic pain management through improved diagnosis and treatment targeting and development.

Individuals from underrepresented racial groups are more likely to suffer from more severe chronic pain, possibly due to chronic stress associated epigenetic mechanisms [20], and financial stress increases vulnerability to pain [21]. Women are also particularly susceptible to the adverse effects of chronic pain [22], and this is underscored by the pain outcome data from the present study population which was predominantly female with a relatively high average pain score. While substantial changes to SES and/or exposure to race related psychosocial stress may be difficult, improvements in how stress is perceived, self-efficacy, and sleep are modifiable clinical targets for lessening pain in at-risk populations.

The current findings support previous reports of poor prediction of average pain in the last week and pain severity compared to pain inference, as well as the development and use of more extensive measures of chronic pain, such as the PROMIS 29 [23 24]. This may be due to the more comprehensive and functional nature of the assessment of interference compared with average pain and pain severity. A prior study investigating chronic pain perception and acceptance found that early changes in pain acceptance were associated with key pain outcomes, yet not associated with pain intensity during the 4-week program [25]. Although changes in depression symptomology may not impact pain severity, they may be a robust indicator of how pain impacts everyday life.

Depression is often strongly correlated with chronic pain, chronic pain can adversely affect the brain, leading to depression [26], and perceived stress is a common contributing factor to both depression and the perception of pain. Chronic low back pain is linked to elevated risks of anxiety and depression, especially in females [27]. Severe depression is associated with total health care costs in chronic pain patients even when controlling for key demographic, functional, and clinical covariates, and effective treatment of depression may improve functional health of patients and attenuate the impact of chronic pain on health care [28]. However, the nature of this relationship in different ethnic groups is unclear, as the reporting of both pain and depression may vary with ethnicity due to differential healthcare related behavior [29]. Nonetheless, our results do support the following mediators as important factors underlying this relationship: perceived stress, sleep quality, and pain self-efficacy.

Severe depression is also associated with higher levels of perceived stress, and the attenuation of stress results in decreased depressive symptoms [30]. Individuals with higher perceived stress levels experience pain more frequently, and pain intensity increases with the perception of stress [31 32]. The significant mediation of the predictive relationship between depression and pain interference by perceived stress in the present study indicates that interventions targeting the attenuation of stress, such as mindfulness-based therapies, may be particularly effective in decreasing pain interference. This also indicates that the assessment of stress can be used as a proxy for depression in similar populations where there may be cultural and/or societal barriers to the reporting and treatment of mood disorders [33 34].

Sleep disturbances are common in both chronic pain and major depression. There is evidence of a direct effect of chronic pain on poor sleep quality, which then adversely affects depression [35]. Chronic pain itself may disturb sleep so extensively that major depression has minimal additive effects [36] and pain intensity has been specifically linked with disordered sleep [37]. Patients with comorbid depression and chronic pain are more likely to express dysfunctional beliefs about sleep and exhibit greater pre-sleep arousal [36]. Individuals suffering from chronic low back pain and major depression or anxiety disorders exhibit greater pain severity and insomnia compared to those without. Major depressive episodes are strongly associated with insomnia, and depression severity may have a stronger association with insomnia than pain intensity [38]. A cross-sectional study investigating the comorbidity of insomnia, chronic pain, and depression suggested that the mesolimbic dopamine system could be a key common neurobiological factor [39]. As with insomnia associated subtypes of depression, the inclusion or targeting of sleep quality in chronic pain patients with disordered sleep may result in the attenuation of the adverse impacts of pain, whether this is directly on pain severity or through improvements in comorbid depression.

Lastly, pain self-efficacy is a key factor in the development of disability and depression in patients with chronic pain [40]. A lack of confidence in being able to manage pain is a significant predictor of chronic pain and associated depression in general [41], and a focal mediator in the present diverse, low-income population. The association between changes in depression and affective pain fully accounted for the change in self efficacy in a study of low back pain [42]. Self-efficacy has been postulated to infer resilience to the adverse outcomes of the depression-pain pathway [43]. Similar to perceived stress, a strong emphasis on self-efficacy in pain management plans and interventions may be particularly effective at reducing pain interference, especially when in conjunction with simultaneous emphases on sleep quality and stress.

Several features of the underlying RCT should be considered in interpreting the outcomes of the present analyses. For example, it is possible that 9 weeks of active in-person group visits was not long enough to see a significant change when comparing routine primary care to medical group visits. In addition, at enrollment, some patients may have had heightened pain and depressive symptoms, and as time went on, their scores may have regressed to the mean. Additionally, the results of this study are based upon subjective responses to self-report measures. Subjective self-report carries known caveats including response biases and inaccurate retrospective accounts of experiences. Suggestions for future research include larger sample sizes, more comprehensive assessment of perceived stress, sleep quality, and/or self-efficacy, and the inclusion of longitudinal manipulative studies to determine causality.

This investigation adds to the considerable literature on the relationship between depression and chronic pain with novel findings from a high-risk, under-represented population. These findings underscore the value of the broad assessment of pain impact. Namely, while depression reliably predicted pain interference, it did not predict average pain or pain severity. Furthermore, mediation analyses identified key mediators in this pathway. Perceived stress, sleep, and pain self-efficacy may be appropriate targets for attenuating the adverse effects of depression on functional outcomes. In the context of the associated RCT and similar interventions, these insights may inform future treatment optimization, targeting, and application efforts. Therefore, this work not only highlights depression as a risk factor for functional impairment, but also psychosocial mediators that may drive these processes.

## Data Availability

Data cannot be shared publicly because of patient confidentiality. Data are available from the Boston Medical Center Institutional Data Access / Ethics Committee (contact via email) for researchers who meet the criteria for access to confidential data.

